# Brain Networks Associated With COVID-19 Risk: Data From 3,662 Participants

**DOI:** 10.1101/2021.04.01.21254709

**Authors:** Chadi G. Abdallah

**Affiliations:** Michael E. DeBakey, VA Medical Center, Houston, TX, USA; Menninger Department of Psychiatry, Baylor College of Medicine, Houston, TX, USA; National Center for PTSD, Clinical Neurosciences Division, US Department of Veterans Affairs, West Haven, CT, USA; Department of Psychiatry, Yale University School of Medicine, New Haven, CT, USA

**Author notes:** **Correspondence to:** Chadi G. Abdallah, M.D.; Menninger Department of Psychiatry, Baylor College of Medicine; 1977 Butler Blvd, E4187, Houston, TX, 77030.

## Abstract

**Background:** Our behavioral traits, and subsequent actions, could affect the risk of exposure to the coronavirus disease of 2019 (COVID-19). The current study aimed to determine whether unique brain networks are associated with the COVID-19 infection risk.

**Methods:** This research was conducted using the UK Biobank Resource. Functional magnetic resonance imaging scans in a cohort of general population (n=3,662) were used to compute the whole-brain functional connectomes. A network-informed machine learning approach was used to identify connectome and nodal fingerprints that are associated with positive COVID-19 status during the pandemic up to February 4^th^, 2021.

**Results:** The predictive models successfully identified 6 fingerprints that were associated with COVID-19 positive, compared to negative status (all *p* values < 0.005). Overall, lower integration across the brain modules and increased segregation, as reflected by internal within module connectivity, were associated with higher infection rates. More specifically, COVID-19 positive status was associated with 1) reduced connectivity between the central executive and ventral salience, as well as between the dorsal salience and default mode networks; 2) increased internal connectivity within the default mode, ventral salience, subcortical and sensorimotor networks; and 3) increased connectivity between the ventral salience, subcortical and sensorimotor networks.

**Conclusion:** Individuals are at increased risk of COVID-19 infections if their brain connectome is consistent with reduced connectivity in the top-down attention and executive networks, along with increased internal connectivity in the introspective and instinctive networks. These identified risk networks could be investigated as target for treatment of illnesses with impulse control deficits.

## Introduction

*“What’s natural is the microbe. All the rest—health, integrity, purity (if you like)—is a product of the human will, of a vigilance that must never falter. The good man, the man who infects hardly anyone, is the man who has the fewest lapses of attention.”* – Albert Camus, The Plague

Chronic stress pathology has been increasingly recognized as a major factor in the pathophysiology of neuropsychiatric disorders ^1^. For certain mental illnesses*—*for example posttraumatic stress and major depression*—*trauma and stress could be the triggering and/or the perpetuating factors. While for others, such as anxiety and paranoia, chronic stress is a detrimental outcome that may exacerbate the underlying pathology ^2, 3^. Furthermore, biological correlates of stress-related disorders may reflect predisposing components and/or an outcome of chronic stress pathology. For example, reduced hippocampal volume is believed to be both a predisposing factor as well as an outcome of posttraumatic stress disorder (PTSD) ^4-6^. Hence, although it is important to determine the brain correlates of chronic stress, it is critical to disentangle the predisposing markers from the consequences of stress. Capitalizing on the fortuitous large neuroimaging dataset from the UK Biobank ^7^, the aim of the current report is to identify the brain signatures that predisposed the general population to a major worldwide stressor, the coronavirus disease of 2019 (COVID-19).

In early 2020, the severe acute respiratory syndrome coronavirus 2 (SARS-CoV-2) infections spread globally instigating a devastating pandemic ^8^. By November 2021, more than 259,000,000 cases were identified, and more than 5,000,000 deaths were related to COVID-19. From life-threatening hospitalizations and the loss of loved ones, to lockdowns, isolation, and increased unemployment and domestic conflict, the impact of the pandemic has been overwhelming ^8, 9^. Moreover, COVID-19 may cause direct damage to the brain through encephalopathy ^10^. Thus, in the ensuing years, it will be essential for the field to assess the long-term impact of the pandemic on mental health and brain function. Equally important is the need to determine the brain signatures that predate the pandemic but correlate with higher SARS-CoV-2 infection rate. Identifying these biomarkers will help us disentangle the sequelae of COVID-19 from its predisposing brain markers. It will also provide a greater understanding of the brain role in the spread of the disease, which may assist in developing future preventive strategies.

This report will focus on the role of the brain intrinsic connectivity networks, using functional connectome fingerprinting. This machine-learning approach allows full assessment of the brain connectome, while providing network informed results ^11, 12^. It is a combination of the network-restricted strength (NRS) ^13^ and connectome-based predictive modeling approaches ^14^. Functional connectome fingerprints (CFPs) were reported to predict behavior in the general population ^15, 16^ and treatment response in depressed patients ^12, 17, 18^. This NRS predictive model (PM) approach has several major strengths. First, predictive features can be back-translated to the original space, which is not often the case in machine-learning algorithms. Thus, instead of establishing a “black box” computational algorithm that is predictive of the outcome but is undiscernible, the NRS-PM works to identify the brain biomarker that is associated, significantly and consistently, with the outcome of interest (e.g., infection status) regardless of the intensity of prediction. Second, the NRS-PM approach could enhance reproducibility by providing protection against overfitting, which is an issue with traditional interpretive statistics. Third, the multivariate pattern analysis permits the full assessment of the connectome, without the inherent increase of Type I error due to univariate multiple comparisons or the need to restrict the analysis to a limited selection of seeds and targets. Finally, the NRS-PM results are network-based by design, which both informs the neurobiological models and facilitates the integration of findings ^17, 19^.

Based on the Akiki-Abdallah (AA) hierarchical connectivity atlas ^11, 12^, the brain connectome is divided into 7 canonical networks: 1) central executive (CE); 2) default mode (DM); 3) ventral salience (VS); 4) dorsal salience (DS); 5) subcortical (SC); 6) sensorimotor (SM); and 7) visual (VI) ^11, 12^. In an environment with multiple priorities and stimuli competing for our attention, two brain systems, the DS and VS, dictate which stimuli is deserving of our attention ^20^. The DS, sometimes called the dorsal attention network, is involved in top-down voluntary attention to salient stimuli. In contrast, the VS network is primarily responsible for reorienting brain resources in response to involuntary salient (i.e., important or conspicuous) external and internal stimuli ^21^. These brain systems interact with the DM and CE networks, which are responsible for internally and externally directed cognitions, respectively ^22^. While the function of the SC network remains unknown, it is hierarchically derived from the salience system and was previously found to complement connectivity changes in the CE ^12, 17^. The current study conducted a data-driven approach assessing all whole brain networks. However, considering the hypothesized role of the brain networks ^23^, it is conceivable to anticipate increased COVID-19 infections in individuals with reduced connectivity in the top-down, attention and executive, control networks (i.e., DS and CE). Here, it is important to note that the goal of the connectome fingerprint modeling in this study is not to establish a classifier risk group stratification, but to conduct an analysis that could identify the brain risk networks underlying the traits vulnerability to SARS-CoV-2 infections.

## Methods

Data used in this study were extracted from the UK Biobank data repository under application number 42826. All study procedures were approved by Institutional Review Boards and all participants completed an informed consent process.

### Participants

The UK Biobank is a prospective epidemiological study of approximately 500,000 participants. Details of the UK Biobank resource and procedures can be found online (https://www.ukbiobank.ac.uk) and in previous reports ^24^. Briefly, between 2006 and 2010, community-dwelling general population individuals across the United Kingdom (n = 502,536; 40 to 69 years of age at the time of recruitment) provided extensive genetic, physical, and health data^24^. In 2016, a followup imaging study was funded to scan 100,000 participants from the existing cohort (including brain, abdomen, heart, and whole-body scans) ^7^.

### Imaging Data

This study used three UK Biobank brain imaging modalities acquired on Siemens Skyra 3T magnet (see ^7, 25^ for more details), including structural high-resolution MRI (T1; 1×1×1 mm), resting-state *f*MRI (2.4×2.4×2.4 mm; 490 frames in 6min.), and task *f*MRI (2.4×2.4×2.4 mm; 332 frames in 4 min.; Hariri faces/shapes “emotion” task ^26^). The study used the UK Biobank preprocessed NIfTI files (see ^7^ for more details). Briefly, only “usable” data (i.e., following manual review and auto quality checks ^25^) were used. For all modalities: quality check scores were generated, based on alignments and signal-to-noise ratios; gradient distortion correction was applied; and nonlinear transformations between native and standard spaces were generated. B0 fieldmaps were used to correct EPI distortion for *f*MRI. In addition, structural MRI preprocessing included tissue-type segmentation using FAST (FMRIB’s Automated Segmentation Tool ^27^) and subcortical structure modeling using FIRST (FMRIB’s Integrated Registration and Segmentation Tool ^28^). Preprocessing of *f*MRI scans included: motion correction, grand-mean intensity normalization, high-pass temporal filtering (sigma=50s), and structured artefact removal by ICA+FIX processing (Independent Component Analysis followed by FMRIB’s ICA-based X-noiseifier ^29^).

### Connectome and Nodal Predictive Models

Full details of the network restricted strength predictive model (NRS-PM) methods were previously reported ^12, 13, 17^ and are described in the Supplements. Briefly, individual specific FAST and FIRST segmentations were used to extract the average time series of 424 nodes that cover the whole-brain gray matter based on the A424 atlas ^12, 30-32^. Full description of the nodes and network affiliations, including A424 projected in the volume space, are publicly available at https://github.com/emergelab/hierarchical-brain-networks/tree/master/brainmaps. The Akiki-Abdallah hierarchical connectivity at 50 modules (AA-50; Fig. S1), 24 modules (AA-24; Fig. S2), and 7 modules (AA-7) were used to determine the network affiliation of the A424 nodes (https://github.com/emergelab). The full connectome is the Fisher-Z transformation of the pairwise correlation coefficients. NRS connectome is the pairwise average connectivity of all modules at AA-50, AA-24 and AA-7 ^12^. Nodal strength (nS) is the average connectivity of a node to all other nodes. Nodal internal NRS (niNRS) is the average connectivity between each node and all other nodes within the same canonical connectivity network (i.e., AA-7). Nodal external NRS (neNRS) is the average connectivity between each node and all other nodes outside its canonical connectivity network ^12^. The predictive models used were adapted from the connectome-based predictive model approach by Shen et al. ^14^, as previously detailed ^12^. All NRS-PM functions used in the current study are publicly available at https://github.com/emergelab. The modeling includes feature selection in training subsamples, followed by fitting a linear predictive model, then applying the model to the test subsample ^14^. Finally, 200 iterations of ten-fold cross-validation (CV) were conducted to ensure the stability of the models and to determine the statistical significance; that is by comparing true and random predictions ^12^. The predictive model included both resting and task *f*MRI connectome data to improve the study predictions ^33^.

### Statistical analyses

Descriptive statistics were calculated prior to statistical analysis. Data distributions were checked using normal probability plots. The statistical significance threshold was set at 0.05 (2-tailed tests). MATLAB (2018a; Mathworks Inc.) and the Statistical Package for the Social Sciences (version 24; IBM) software were used for the analyses. False Discovery Rate (FDR; *q* < 0.05) was used to correct for multiple comparisons. The connectivity fingerprints (CFPs) were examined at AA-50, AA-24, and AA-7. The nodal fingerprints (NFPs) were determined for nS, niNRS, and neNRS. FDR was applied on all 6 outcome measures to determine statistical significance.

As in previous reports ^12, 17^, connectivity per fingerprint was computed by multiplying the connectivity features (e.g., NRS at AA-50) by the corresponding weighted fingerprint masks (e.g., the COVID-19 CFP at AA-50). Thus, the CFP total connectivity is the sum of weighted estimates per subject per CFP. To facilitate the comparison across measures, the CFP connectivity values were standardized (z-scored). Follow-up analyses covarying for age and sex used general linear models with the 6 fingerprints’ total connectivity as dependent variables and COVID-19 status as fixed factor. The study atlases, code, and predictive models will be made publicly available at https://github.com/emergelab.

## Results

The brain imaging data were based on a package downloaded on July 8^th^, 2020. At the time the data were downloaded, preprocessed brain imaging data from 40,681 participants were available for this report. The COVID-19 results were downloaded on February 4^th^, 2021. The COVID-19 data included 60,446 UK Biobank participants, of which 3,662 had successful structural MRI, and resting and task *f*MRI scans. These 3,662 individuals were investigated in the current report. They were 52% females (n=1896). Their average age was 63 years (*SEM*=0.13) at the time of the brain scan and 66 years (*SEM*=0.13) at the time of the SARS-CoV-2 testing. A total of 921 (25%) tested positive for SARS-CoV-2. All imaging data used in the current study were acquired prior to the COVID-19 pandemic.

The predictive models successfully identified 6 fingerprints that were significantly associated with COVID-19 positive, compared to negative status: (1) AA-50 CFP (*r* = 0.13, *CV* = 10, *iterations* = 200, *p* < 0.005, *q* < 0.05; Fig. 1A); (2) AA-24 CFP (*r* = 0.13, *CV* = 10, *iterations* = 200, *p* < 0.005, *q* < 0.05; Fig. 1B); (3) AA-7 CFP (*r* = 0.11, *CV* = 10, *iterations* = 200, *p* < 0.005, *q* < 0.05; Fig. 1C); (4) nS NFP (*r* = 0.09, *CV* = 10, *iterations* = 200, *p* < 0.005, *q* < 0.05; Fig. 2A-B); (5) neNRS NFP (*r* = 0.10, *CV* = 10, *iterations* = 200, *p* < 0.005, *q* < 0.05, Fig. 2C); and (6) niNRS NFP (*r* = 0.12, *CV* = 10, *iterations* = 200, *p* < 0.005, *q* < 0.05; Fig. 2D).

**Figure 1.**
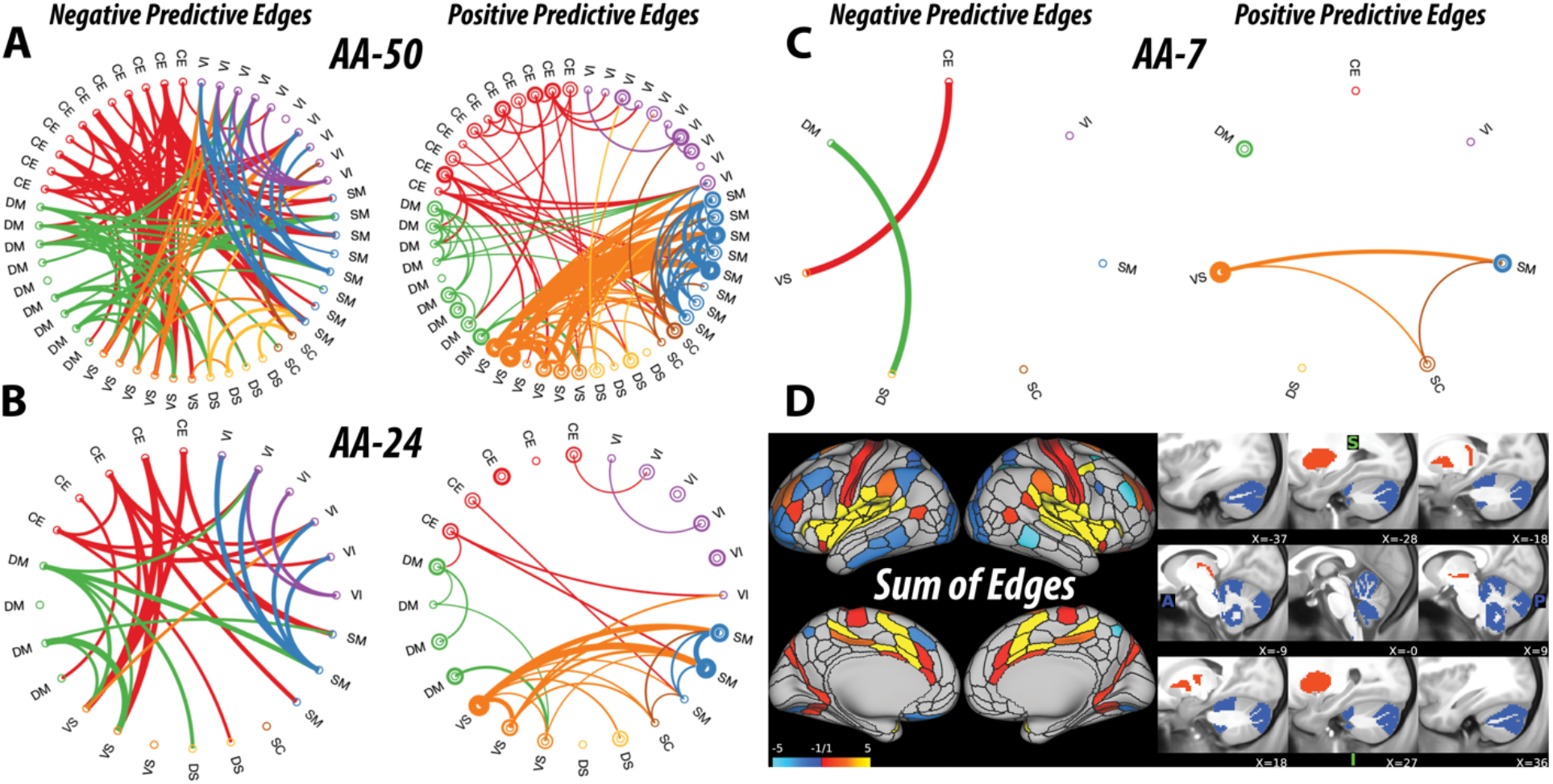
COVID-19 Connectome Fingerprint (CFP). **A-D**. Predictive models applied to functional magnetic resonance imaging scans, acquired years before the pandemic in a general population cohort of older adults, identified unique CFPs that predict higher COVID-19 infection in individuals with reduced connectivity between the brain networks (see the Negative Predictive Edges) but increased internal connectivity within networks and their underlying modules (see the Positive Predictive Edges). *Notes*: The circular graphs are labeled based on the Akiki-Abdallah (AA) whole-brain architecture at 50 modules (AA-50), 24 modules (AA-24), and 7 modules (AA-7). Modules and nodes are colored according to their affiliation to the 7 canonical connectivity networks: central executive (CE), default mode (DM), ventral salience (VS), dorsal salience (DS), subcortical (SC), sensorimotor (SM), and visual (VI). Edges are colored based on the initiating module using a counterclockwise path starting at 12 o’clock. Internal edges (i.e., within module) are depicted as outer circles around the corresponding module. Thickness of edges reflect their corresponding weight in the predictive model. The module abbreviations of AA-7, AA-24 and AA-50, along with further details about the affiliation of each node are available at https://github.com/emergelab/hierarchical-brain-networks/blob/master/brainmaps/AA-AAc_main_maps.csv. Only edges of significant predictive models following correction are shown (all *p* < 0.005). Panel D shows the nodal degree of the AA-50 fingerprint edges. The color bar unit is arbitrary, reflecting the sum of weighted edges. All predictive models will be made publicly available at https://github.com/emergelab.

**Figure 2.**
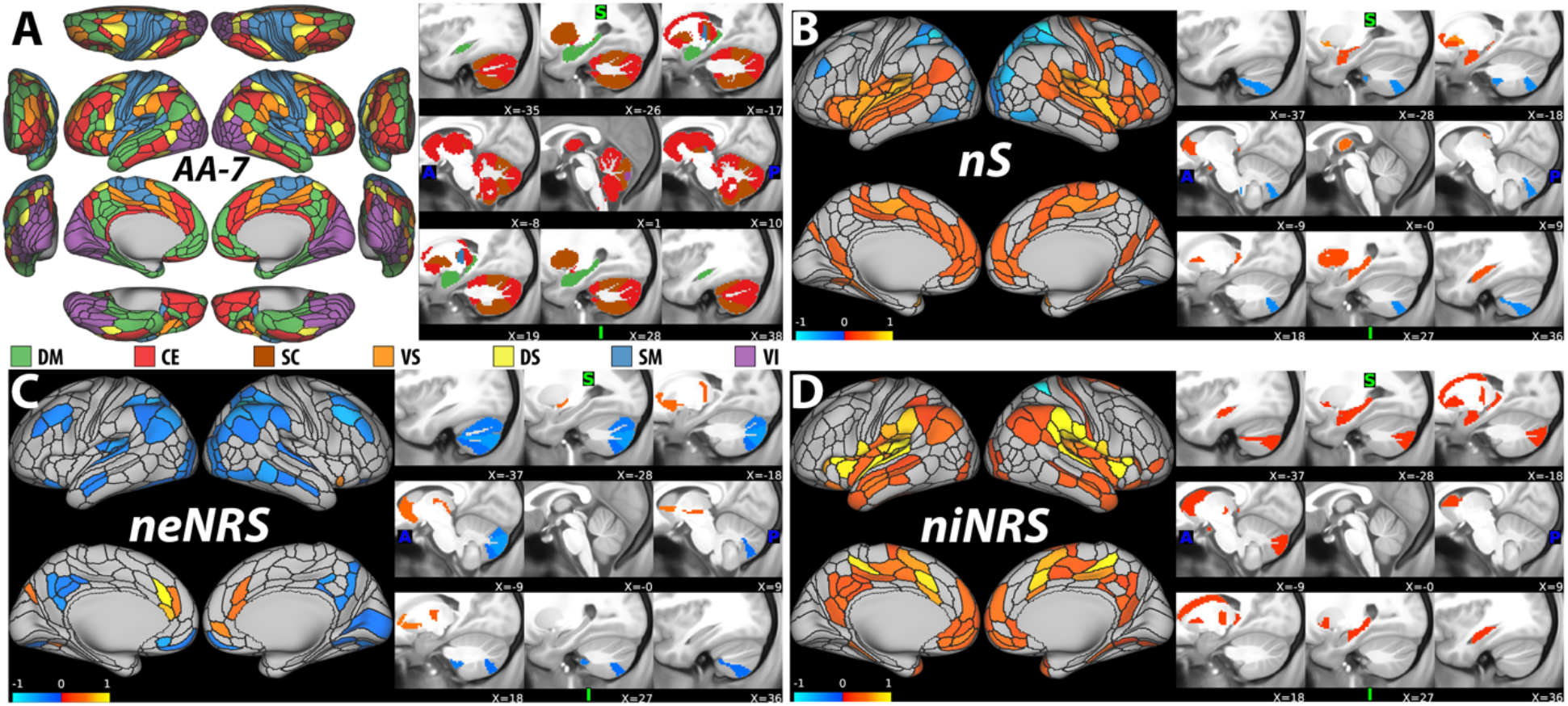
COVID-19 Nodal Fingerprint (NFP). **A**. The nodal affiliation based on the Akiki-Abdallah (AA) hierarchical atlas at 7 canonical intrinsic connectivity networks (i.e., AA-7): default mode (DM), central executive (CE), subcortical (SC), ventral salience (VS), dorsal salience (DS), sensorimotor (SM) and visual (VI). The AA-7 affiliation was used to compute nodal external network restricted strength (neNRS) and nodal internal NRS (niNRS). **B-D**. Nodal predictive results using nodal strength (nS; **B**), neNRS (**C**), or niNRS (**D**) as input features in general population older adults tested for COVID-19 infection status. The nS findings (**B**) associated positive COVID-19 status with increased global connectivity in the VS and DM (red-yellow), but reduced connectivity in DS and CE networks (blue). The neNRS (**C**) and niNRS (**D**) findings demonstrate a connectivity shift with increased internal within network connectivity in the VS and DM but reduced external connectivity in the DS and CE. *Notes*: Only nodes of significant predictive models following correction are shown (all *p* < 0.005). The color bar unit is arbitrary, reflecting the sum of weighted nodes. All predictive models will be made publicly available at https://github.com/emergelab.

As shown in Fig. 1, positive COVID-19 tests were associated with increased internal connectivity within modules but reduced external connectivity between the brain networks. In particular, positive COVID-19 tests were associated with reduced connections between the central executive (CE) and ventral salience (VS), as well as between the dorsal salience (DS) and default mode (DM) modules. In contrast, increased interference from the VS to the sensorimotor (SM) and subcortical (SC) networks were associated with positive COVID-19 results (Fig. 1C).

The external to internal connectivity shifts observed in the CFPs were translated into overall reduced neNRS but increased niNRS as shown in Fig. 2. Independent of network constraints, positive COVID-19 results were associated with increased overall functional connectivity strength (i.e., nS) in the insula and surrounding regions, as well as in the medial frontal area (Fig. 2B).

To account for the effect of age, a general linear model examined the effects of COVID-19 status on the 6 fingerprints’ total connectivity covarying for age (Fig. 3). This multivariate test showed statistically significant effects of COVID-19 status on the fingerprints’ connectivity (F = 2.7, *p* = 0.01, *q* < 0.05). Post-hoc univariate tests of between-subject effects were significant for nS (F = 14.3, *p* < 0.001, *q* < 0.05), niNRS (F = 10.8, *p* = 0.001, *q* < 0.05), and neNRS (F = 9.6, *p* = 0.002, *q* < 0.05), but not for CFPs at AA-50 (F = 3.5, *p* = 0.060, *q* > 0.05), AA-24 (F = 3.6, *p* = 0.059, *q* > 0.05), and AA-7 (F = 1.0, *p* = 0.31, *q* > 0.05). Covarying for sex did not affect the main study results with all 6 fingerprints retaining significance (all *p* values < 0.001, *q* < 0.05).

**Figure 3.**
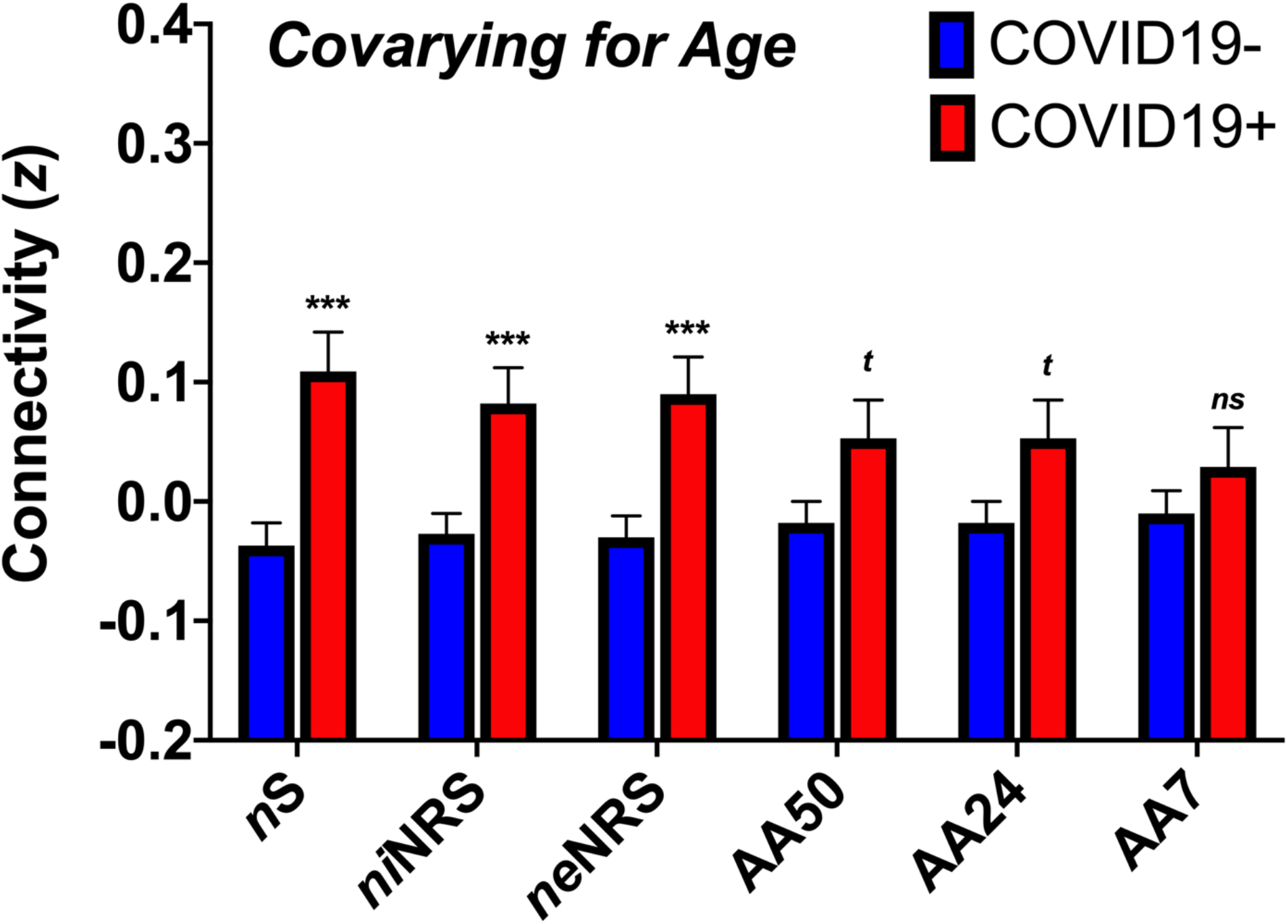
COVID-19 Fingerprints Covarying for Age. The COVID-19 functional connectivity fingerprints were examined covarying for age. Positive COVID-19 status remained significantly associated with the nodal strength (nS), nodal internal network-restricted strength (niNRS) and nodal external NRS (neNRS) fingerprints. However, COVID-19 status was associated only at a trend level with the connectome fingerprints (CFPs) of Akiki-Abdallah (AA) hierarchical connectivity atlas at 50 modules (AA50) and 24 modules (AA24) but not 7 modules (AA7). *<u>Abbreviations</u>* – *** is used for *p* ≤ 0.001, ***t*** for *p* ≤ 0.1. *z* is computed as the standardized sum of weighted estimates of connectivity per subject per fingerprint.

## Discussion

This report successfully identified brain functional connectivity risk networks that were significantly associated with SARS-CoV-2 status in a relatively large general population sample of older adults. The results showed that individuals with positive status tended to have lower integration across the brain connectivity networks but increased internal connectivity within modules. Together, these findings indicate that a shift toward increased segregation between the brain networks is a putative marker of increased risk-taking traits. SARS-CoV-2 infections were associated with reduced connectivity between the top-down, attention (DS) and executive (CE) control networks and the introspective (DM) and instinctive (VS) networks, respectively. Importantly, the DS-DM and CE-VS reductions in top-down connectivity were accompanied by increased internal connectivity in both the DM and VS networks. This shift from external to internal connectivity was evidenced in the frontoparietal reduction in neNRS and increased niNRS in the insular and medial frontal brain regions. At the level of global connectivity as measured by nS, SARS-CoV-2 infections were associated with increased connectivity in areas within the VS and DM networks.

Considering that the study scans were acquired an average of 3 years prior to the COVID-19 testing, the results suggest that these identified risk networks may reflect personality traits of increased impulsivity or risk-taking phenotypes. Notably, the *f*MRI data cannot directly predict SARS-CoV-2 infection, but it could be putatively detecting a network that is associated with behavioral traits that are in turn related to an increased chance of exposure to the virus. For example, accumulating literature demonstrates the utility of functional brain connectivity in predicting behavioral characteristic of addiction, which may also be related to reduced control and increased risk taking ^34, 35^. Furthermore, it is important to underscore the similarity between the identified risk networks and the brain circuits currently targeted by transcranial magnetic stimulation (TMS) for the treatment of depression ^36-38^. Connectome wide studies have found increased top-down control, as reflected by increased external connectivity in the central executive network, results in enhanced antidepressant response ^12, 17^. Hence, it is plausible that the risk networks identified in this report could be successfully targeted for depression treatment by TMS or other modalities.

Considering the scope and resources of the current report, a limitation of the study is the lack of full assessment of behavioral measures available through the UK Biobank. Investigating the behavioral correlates of the COVID-19 CFPs and NFPs in future studies will be essential to better understand the role of the brain in mitigating infection risk and perhaps develop new approaches to limit the spread of infections in future epidemics. Another limitation is that the causal relationship between the network alterations and infection status cannot be established in this associative study. However, the longitudinal design rules out the possibility that the identified fingerprints are the consequence of SARS-CoV-2 infections. These brain biomarkers could be an underlying factor to increasing infection risk. They may also be a confound or compensatory response to an unknown underlying causal factor. While the COVID-19 NFPs retained significance when controlling for age and sex, the study failed to rule out the possibility that the variance in the COVID-19 CFPs may be at least partially affected by age as a confound. Another limitation is that the study cannot determine whether the results will generalize to younger adults. Future studies should further investigate the role of age in the brain risk networks.

The study has several strengths, including: 1) large cohort; 2) longitudinal data; 3) high quality imaging acquisition and preprocessing using standardized methods; 4) data-driven approach that is not limited to a biased selection of seeds; 5) network informed design to facilitate the interpretation and guide future follow-up studies; and 6) urgently needed data to better understand a devastating pandemic and ultimately devise additional preventive measures to improve global health and reduce suffering. It is rare to have such a large neuroimaging sample collected prior to unpredictable traumatic stressors like disasters or epidemics. Not only does this allow future studies of post-pandemic neural alterations to disentangle consequence of disease and chronic stress from predisposing characteristics, but it also underscores the tremendous value of large, prospective, longitudinal, multi-modal epidemiological efforts like the UK Biobank, and the critical need to continue funding similar projects that cover the life-span and other geographic regions.

## Data Availability

Data are publicly available through the UK Biobank.

https://bbams.ndph.ox.ac.uk/ams/

## Acknowledgement

This research has been conducted using the UK Biobank Resource under Application Number 42826. I would like to thank all the UK Biobank investigators, staff and participants for contributing to this tremendous effort. I also would like to thank the Emerge Research Program (<u>emerge.care</u>) staff and investigators, who made possible the line of research conducted in this study.

## Funding

This work was supported by the Department of Veterans Affairs, the VA National Center for PTSD and the Beth K and Stuart Yudofsky Chair in the Neuropsychiatry of Military Post Traumatic Stress Syndrome.

The funding sources have had no involvement in the study design, the analysis and interpretation of data, the writing of this report, or the decision to submit this article for publication.

## Declaration of Conflicting Interests

Dr. Abdallah has served as a consultant, speaker and/or on advisory boards for Genentech, Janssen, Psilocybin Labs, Lundbeck, Guidepoint, and FSV7, and as editor of *Chronic Stress* for Sage Publications, Inc. He also filed a patent for using mTORC1 inhibitors to augment the effects of antidepressants (Aug 20, 2018).

## SUPPLEMENTS

**Figure S1.**
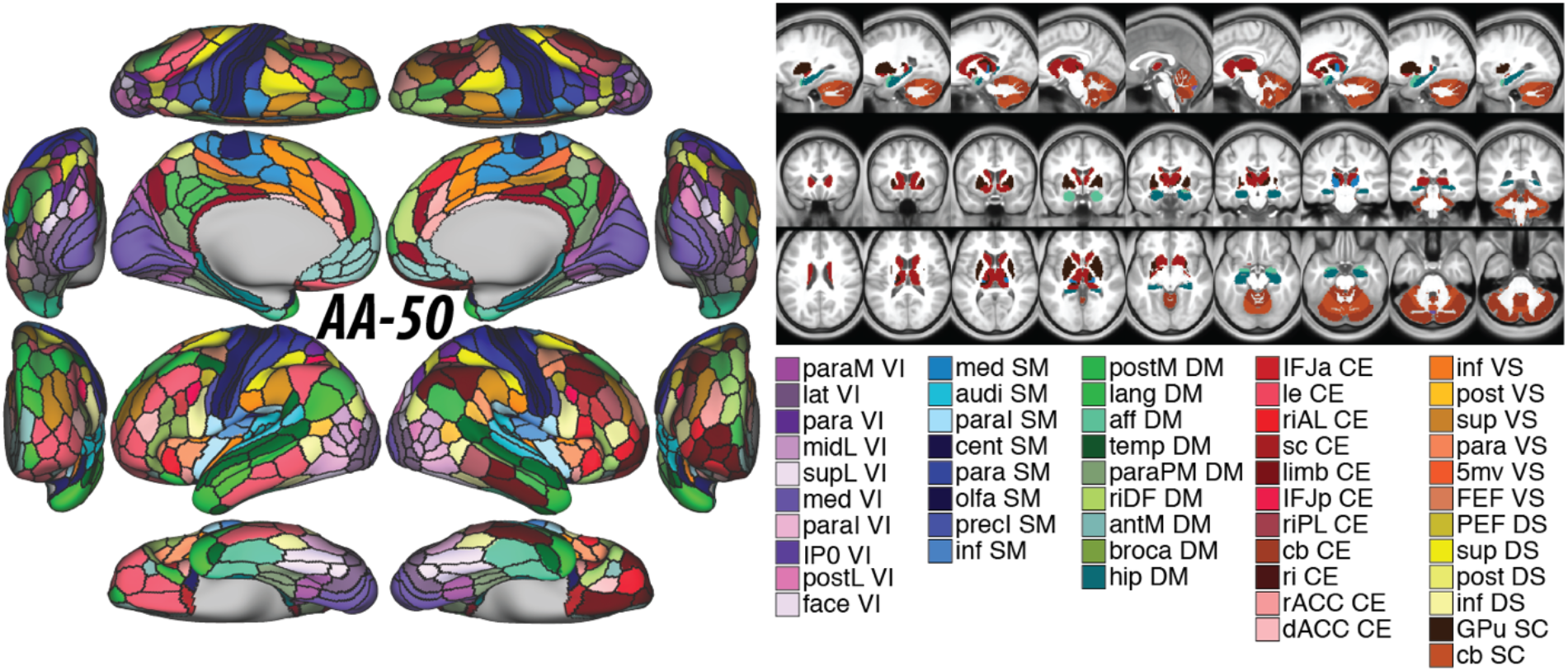
Whole-brain Akiki-Abdallah (AA) network affiliation at 50 modules architecture (i.e., AA-50). The module abbreviations of AA-50, along with further details about the affiliation of each node are publicly available at ^1^ and at https://github.com/emergelab/hierarchical-brain-networks/tree/master/brainmaps. The figure was adapted with permission from the Emerge Research Program (http://emerge.care).

**Figure S2.**
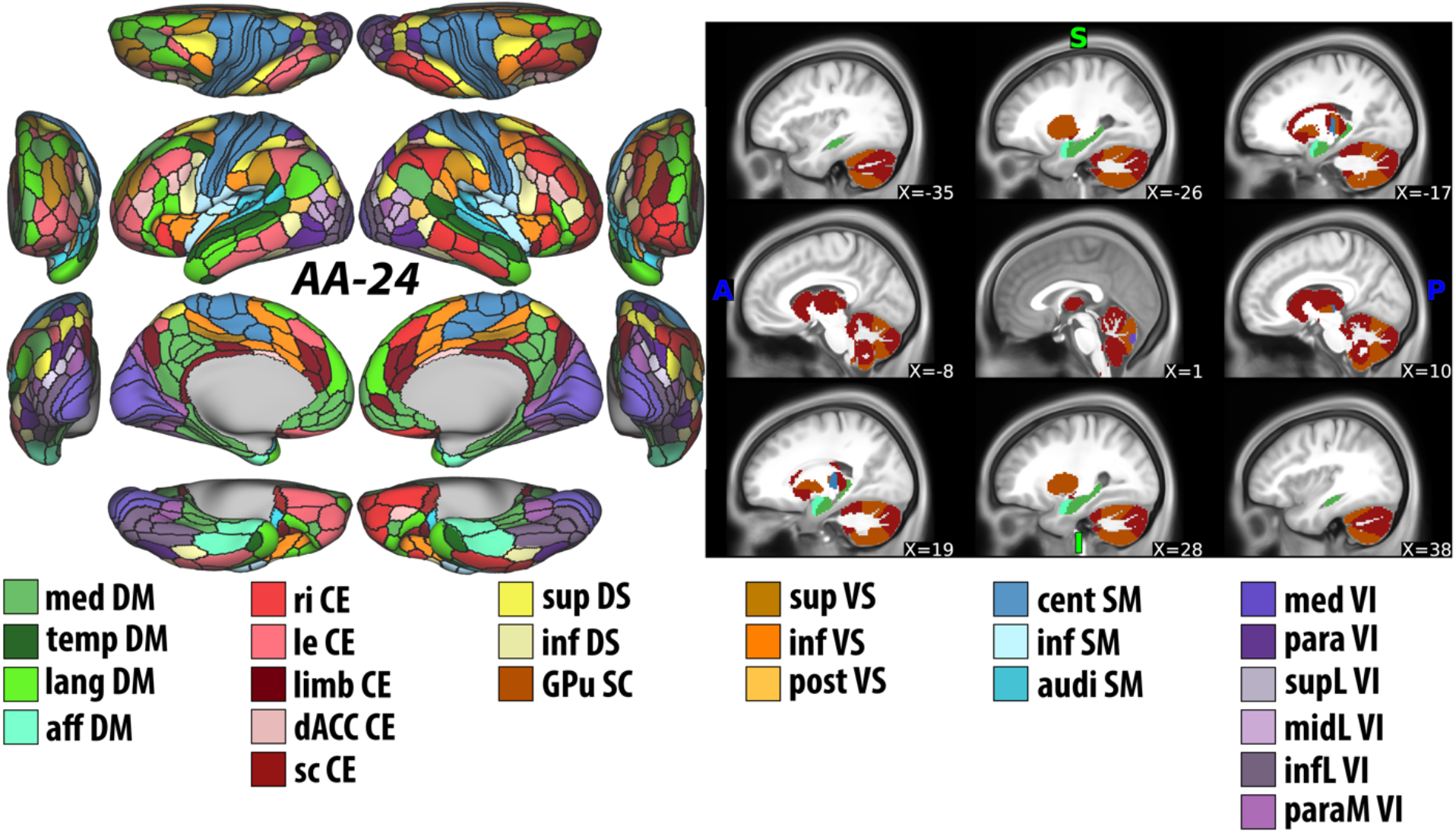
Whole-brain Akiki-Abdallah (AA) network affiliation at 24 modules architecture (i.e., AA-24). The module abbreviations of AA-24, along with further details about the affiliation of each node are publicly available at ^1^ and at https://github.com/emergelab/hierarchical-brain-networks/tree/master/brainmaps. The figure was adapted with permission from the Emerge Research Program (http://emerge.care).

### Nodal Parcellation and Network Restriction

The brain nodes were defined using previously established multimodal parcellation atlases that divide the cerebral cortex ^2^, subcortical regions ^3^, and the cerebellum ^4^ into 424 nodes within the grayordinate, here termed A424 ^1^. Detailed maps and nodal affiliations can be found at https://github.com/emergelab/hierarchical-brain-networks/tree/master/brainmaps. Within each node, an averaged time series of all voxels/vertices was calculated. The full connectome was computed as the pairwise Pearson correlation coefficients between these averaged time series, and subsequently transformed using a Fisher-z function.

To define the network modules, we used the Akiki-Abdallah (AA) atlas ^1, 5^, a recently described hierarchical modular organization of the brain based on the same parcellation atlas used in the current study. The AA atlas provided an extensive characterization of the whole-brain modules at multiple scales, ranging from three communities at the first hierarchical split to 150 communities at the finest-grained level.

To test the hypothesis that the connectivity in the canonical networks can predict risk, we used the AA architecture at 7, 24 and 50 modules, which were previously identified as key architectural levels ^1, 6^.

### Predictive Modeling

Traditional interpretative analyses are informative in that they can highlight links between brain and behavior. However, they may not be adequate to predict outcomes and could suffer from over-fitting ^7^. The connectome predictive model (CPM) described by Shen et al. ^8^, takes full connectome matrices as inputs and its utility has been well validated in multiple studies (e.g., ^9^). Building on the strengths of CPM, comparable approaches were previously implemented: 1) termed network-restricted strength predictive model (NRS-PM); 2) termed nodal strength predictive model (nS PM); 3) termed nodal external NRS-PM (neNRS-PM), and 4) termed nodal internal NRS-PM (niNRS-PM) ^1, 6^.

The NRS-PM builds a predictive model of the behavioral data based on NRS edges (i.e., internal and external connectivity). The first step serves as feature selection: the within- and between-network connectivity values (i.e., edges) from the training subset of the data (described below) are used in regression models (Pearson correlation) to identify NRS edges that positively or negatively predict the outcome measure of interest (*p* < 0.05). The weighted sum of positive edges minus the weighted sum of negative edges generates a summary statistic for each subject that is then included in a linear predictive model ^8^. Next, as described by Shen et al. ^8^, the resulting coefficients are applied to the test dataset to predict the outcome. The correlations between the predicted and actual outcomes (*r* values) are used to judge the model’s performance ^8^. To ensure the stability of the models and to determine the significance of the predictive power, 200 iterations of ten-fold cross-validation (CV) are used, which splits the original sample into 10 random equal sized subsamples. In each iteration, one subsample (10% of subjects) is retained as the validation data for testing the model, and the remaining 9 subsamples (90% of subjects) are used as training data. The cross-validation process is then repeated 10 times (covering all subsamples). Then, the predicted outcome is separately correlated with the true outcome (i.e., true prediction) and with a random permutation of the true outcome (i.e., random prediction). This process is repeated 200 times to generate empirical true and null distributions of the test statistics (i.e., *r*). The *p* values are then computed as the proportion of random predictions that are equal to or greater than the average of true predictions.

The nS-PM, neNRS-PM, and niNRS-PM used the same approach described above for the NRS-PM, with the only difference is in the dimensionality reduction step. For nS-PM, the dimensionality reduction step was computing the average correlation between each node and all other nodes in the full connectome, which generates 424 features. For neNRS-PM, the dimensionality reduction step was computing the average correlation between each node and all other nodes outside its canonical connectivity network (i.e., AA-7), which generates 424 features. For niNRS-PM, the dimensionality reduction step was computing the average correlation between each node and all other nodes within the same canonical connectivity network, which generates 424 features.

## Notes

### Author Declarations

Data used in this study were extracted from the UK Biobank data repository under application number 42826. All study procedures were approved by the UK Biobank Institutional Review Boards and all participants completed an informed consent process.

